# Level of Compliance and Predictors with Personal COVID-19-preventive measures Among Office Government Employees Returning to work in the post-epidemic period in Western Ethiopia: A Multicenter Cross-sectional Study

**DOI:** 10.1101/2022.07.26.22278056

**Authors:** Gebisa Guyasa Kabito, Meskele Abreham, Amensisa Hailu Tesfaye, Tadesse Guadu

## Abstract

**Background:** The contemporary global issues, COVID-19 pandemic continued causing unprecedented impact on the public health, occupational health and the global economy. Keeping compliance with personal preventive measures is a vital tool for managing COVID-19 pandemic control and returning to work as no pharmaceutical treatments are currently available in Ethiopia. Although compliance with COVID -19 personal preventive measures (CPPMs) and predictors is well addressed in healthcare settings, data on the level of CPPMs and its determinants among government employees working in offices worldwide, including Ethiopia, is limited. This paper is aimed to fill this gaps.

**Methods:** We applied a cross-sectional study design from February to March, 2021. The participants were government workers working in offices who had resumed work. Stratified followed by simple random sampling technique was used to select 422 study participants from 30 government offices that had resumed work. Data were collected using a pre-tested and structured interviewer-administered questionnaires and analyzed by STATA 14 software. The significance of associations was established at p< 0.05 and adjusted odds ratio (AOR) with 95% confidence intervals (CI) in the multivariable model.

**Results:** Response rate 95.44% (N=394). The study found 22.3% (88) of study participants (95% CI = 18.5, 26.6) had high compliance with COVID -19 personal preventive measures during past month. Female workers were 2.80 times more likely than males to comply with COVID-19 personal preventive measures (AOR: 2.80, 95%CI (1.10, 7.12), favorable attitude towards COVID-19 prevention measures (AOR: 13.73, 95% CI (4.85, 38.83), high-risk perception of COVID-19 infections (AOR: 2.34; 95% CI (1.24, 4.41), and high misconception about COVID-19 (AOR : 3.92, 95% CI (1.45, 10.62) were predicted better compliance with COVID-19 PPMs (P < 0.05).

**Conclusions:** In sum, little proportion of sampled study participants complied with COVID -19 personal preventive measures. Sex, attitudes, risk perception, and misconception have all been identified as significant risk factors. Actions are needed to strengthen COVID -19 personal preventive measures among government employees to maintain COVID -19 control following work resumption. In the future, it’s vital to work on government employees’ attitudes and perceptions in order to improve compliance.

## Introduction

The contemporary global issues, COVID-19 pandemic, has disrupted life and work habits and has produced landmark changes worldwide [1-3]. In light of this, offices, like other workplaces, are a source of stress for both employees and customers, but the nature and extent of work-related stressors in non-healthcare settings have changed dramatically in recent years[4-7]. The pandemic is continuing to have an unprecedented impact on the public health, occupational health and the global economy. As an example, in USA, 644 workers were contracted COVID-19 due occupational exposure at offices, although the actual numbers are unknown because of inadequate data collection systems in the rest of the world[8]. Furthermore, occupational exposure to COVID-19, also been linked to long-term depression, anxiety, and insomnia [9]. As of October 8, 2021, WHO had received reports of 236,599,025 confirmed cases of COVID-19, with 4,831,486 deaths, of which 352,504 were in Ethiopia, with 5,888 deaths [10]. Likewise, as of 19^th^ June 2022, the total number of confirmed COVID-19 cases in Ethiopia has reached 484,138 with 7,523 deaths, 458,280 recovered [11], with Addis Ababa and Oromia reginal state accounting for 65% and 14% of the cases respectively [12, 13]. Indeed, the COVID-19 pandemic has disrupted the global economy, the health systems, social and the quality of life predominantly in developing countries [14, 15]. The global GDP was estimated a 4.5% drop in economic growth results in almost 2.96 trillion U.S. dollars of lost economic output[16].

Global efforts to mitigate the effects of the pandemic and reduce the health and socioeconomic impact rely heavily on personal preventive measures [17]. To slow the spread of the virus, the government initially imposed a nationwide lockdown. However, such restrictions harmed the country’s economy. Ethiopia, as a low-income country, could not afford a prolonged lockdown in the face of unprecedented challenges. As a result, the Ethiopian government decided to ease the lockdown. Although the COVID-19 pandemic has been identified as a major occupational health problem, particularly in healthcare workplace settings, very little research has been conducted to evaluate compliance to personal COVID-19 preventive measures (PPMs) among government office workers globally. Given that, COVID-19 infection can spread quickly among officials, personnel, and visitors at offices and other workplaces with shared spaces such as corridors, elevators & stairs, parking places, cafeteria, meeting rooms and conference halls, and so on. Thus, in order to limit the spread of infection, it is necessary to prevent infection in non-healthcare workplace settings [4].

Similarly, according to WHO, indoor locations, particularly those that are enclosed and confined with little or no ventilation, are riskier than outdoor locations to acquire COVID-19 virus infections[18]. To reduce the risk of infection, it is critical to ensure compliance with personal preventive measures (PPMs) in the government offices. Furthermore, government employees spend an average of 8 hours each day at the workplace, which may be in crowded [5]. Keeping compliance with personal preventive measures is critical to striking a balance between COVID-19 pandemic control and resumed work. The usefulness of these PPMs is heavily reliant on government employees’ compliance at work[19]. Epidemiological studies conducted in China[20], Australia[21], and Thailand[22] consistently supported the importance of achieving high compliance with personal preventive measures (PPMs), ranging from 80% to 95%, in controlling the COVID-19 pandemic.

Personal Preventive Measures (PPMs) against COVID-19 include frequent handwashing with soap and water, physical distancing, wearing facemasks, including homemade masks, using alcohol-based sanitizers, covering mouth and nose while coughing and sneezing, avoiding touching the nose, mouth, and eyes with hands, and refraining from risky behavior, all of which are recommended interventions and heavily promoted and implemented in Ethiopia to control the rate of transmission[23].

The compliance with PPMs is dependent on many factors; for example the existing literatures showed socio-demographic attributes (e.g. being female[24]), perceived effectiveness of preventive measures by the individual[25], media exposure [26], knowledge[27], risk perception[25] were statistically significant with compliance to PPMs.

Therefore, to the best of our knowledge, very limited study has investigated compliance with personal preventive measures and associated factors among government employees who resumed work during the COVID-19 pandemic at office setting in Ethiopia. In the current study, we examined the level of compliance with personal preventive measures against COVID-19 and associated factors among government employees working in offices in Nekemte city administration in western Ethiopia, at the beginning of work resumption.

## Materials and Methods

### Study design and period

An institution based cross-sectional study was conducted from February to March, 2021.

### Study area

The study was carried out in Nekemte town, East Wollega zone, west Ethiopia, which is 335 Km west of Addis Ababa, capital city. It is the center of Western Ethiopia serving as a transient point for different zones and three regional states of the country. Total population of the town is 127,380 among which male constitutes 51.03% [28]. There is one specialized hospital, one referral hospital, two health centers, and seven health posts in the town. Along with 30 governmental organizations/offices in the town with estimated 2500 government employees (civil servant).

### Source and study populations

All government employees (GE) working in offices were the source population. The randomly selected employees in the selected government offices and work in offices were our study population. All government employees who were available in the office was included in the study and those who were unable to communicate easily during data collection were excluded.

### Sample size determination and sampling procedures

The sample size was calculated using a single population proportion formula [29] with a 95% confidence interval (CI), 5% margin of error (*d*), proportion (*P*) 50%, since this would yield the maximum possible sample size. Considering 10% non-response rate the final sample size was 422. To recruit eligible samples, we employed stratified followed by simple random sampling. The list of government employees was obtained from the human resources (HR) departments of each government office in town. So then, a numbered list of all employees in town was created, and study participants were chosen at random using random number generator software. Accordingly, the sample size was proportionally assigned to each government office based on the number of employees in the respective government offices. Participants were approached at their workplaces, using all of the WHO-recommended COVID-19 preventive measures, such as physical distance, face masks, and hand hygiene.

### Data collection tools

Data were collected using a pre-tested and structured interviewer-administered questionnaires methods. The questionnaire was adapted and modified in local settings based on previously available studies [30-32], WHO guidelines[33], and Center of Diseases Control tools[34]. The tool includes seven components: Sociodemographic information, compliance with COVID-19 personal preventative measures, knowledge of COVID-19 and preventative measures, attitudes toward COVID-19 and preventive measures, risk perception of COVID-19 infection, misconceptions, and individual behavioral characteristics. We looked at the Cronbach’s alpha coefficients of each component item, and the results show that they are all satisfactory, ranging from 0.68 to 0.84 across all components [35].

### Operational definitions

Compliance with personal preventive measures against COVID-19 was operationalized as an implementation of the four key COVID-19 preventive measures at workplaces in the past month[36]. These are wearing a face mask, frequent hand hygiene, maintain social distance and respiratory etiquette. Accordingly, participants who used the four key components/ 100% were considered “high compliance” with personal COVID-19 personal preventive measures, otherwise “low compliance”

COVID-19 knowledge was assessed using 13 items adapted from previous research [37]. Items 1-4, 5-8, and 9–13 in this list pertain to participant understanding about COVID-19 clinical manifestations, transmission pathways, and prevention and control, respectively. Study subjects were given “true,” “false,” or “not sure” response options to these items. An item’s accurate response assigned 1 points, whereas an incorrect or unsure response received 0. Respondents were classified as having good knowledge if they scored a median or higher score; otherwise, poor knowledge. Attitude towards COVID-19 and Preventive measures were assessed using 11 questions, with 7 questions about COVID-19 and 4 questions about preventive measures on a five-point ordinal scale “strongly disagree” to “strongly agree”. Participants who responds median and above score of the attitude questions about the COVID-19 were labeled as having favorable attitude otherwise unfavorable attitude. Risk perception about COVID-19 infection was measured with two dimensions, first dimension was symbolize how likely one considered oneself (his/her families) would be infected with COVID-19 if no personal preventive measure will be taken. The second dimension was denote how one rated the seriousness of symptoms caused by COVID-19, their perceived chance of having COVID-19 cured and that of survival if infected with COVID-

19. To determine the respondents’ levels of risk perception, we pooled the two dimensions and asked five items with five response options. Answers were scored on a five-point ordinal scale reflecting the levels of contacting COVID-19, such as “How likely you will be infected? How likely your family will be infected?” Every item on a scale from 1 to 5, ranges in the susceptibility from ‘very unlikely’ to ‘very likely’. Accordingly, response were classified into low perceived risk (“very unlikely” or “unlikely”), (“very low” or “low”), and (“not serious at all” or “not serious”) and high perceived risk (“very likely” or “likely”) [38]. We examined misconception of COVID-19 with 11 questions. Each question had five possible response categories ranging from strongly disagree (1) to strongly agree (5). Hence, respondents who score =or > median were coded as having misconceptions; otherwise, no misconceptions[39]. Furthermore, a number of demographic and behavioral information were also collected, such as age, gender, marital status, pay, employment history, family size, educational attainment, religion, cigarette smoking, khat chewing, alcohol drinking, and physical exercises. Finally, in the current study, when we refer to “government employees,” we only mean those who are employed in office by a government department, agency, or public sector organization.

### Data quality control

The tool was initially prepared in English and then translated into the local language, *Afaan oromo* and back translated to English in order to check its consistency. We recruited ten data collectors and four supervisors with different health-related professional backgrounds. One day training was offered for data collectors and supervisors on topics related to research objectives, clarity of questions, utilization of PPE, the confidentiality of information and consent in the study. The training was given in lecture, roleplay and discussion ways. The questionnaires were pre-tested on 20 samples that were not included in the final analysis and the relevant modifications were made before the actual data collection was conducted.

### Data management and statistical analysis

The data were checked for completeness and entered into Epidata version 4.6, and exported to STATA 14 windows for analysis. Frequency distributions, percentages, means, and standard deviations were used for description of the results. Binary logistic regression (Bivariable and multivariable binary logistic regression) was performed to identify statistically significant variables. Adjusted odds ratio with 95% confidence interval was used to declare statistically significant variables on the basis of p<0.05 in the multivariable binary logistic regression model. Hosmer and Lemeshow goodness-of-fit test was used to check the model fitness (P>0.05).

### Ethics approval and consent to participate

Ethical clearance was obtained from the Institutional Review Board (IRB) of University of Gondar (Reference #: IPH/625/2021) and an official permission letter was gained from the Nekemte city administrative office. Written informed consent was obtained from each participant before conducting the actual data collection process for guaranteeing their choice of participation or refusal. Any identifiable issues were eliminated to ascertain confidentiality. Furthermore, an appropriate COVID-19 infection prevention measures were considered during data collection process.

## Results

### Socio-demographic and behavioral characteristics of study participants

A total of 394 questionnaires were completed making a response rate of 95.44%. The mean (±SD) age of the participant was 38.39 (± 6.80) years, accompanied by most (42.4%) of their age group found between 36-44 years. More than half, (53.3%) of the participants were females and three-fourths, (74.6%) of the participants were married. Regarding educational status, 18.8%, 19.8%, and 61.4% of the participants had certificates, diplomas, and degree receptacles respectively. One-fourth (25.1%) of participants had indicated 5-10 years of working experience. Concerning, monthly salary 3 out of 7 (42.9%) participants’ wage was found between 5201-7200 Ethiopian Birr (ETB). Regarding behavioral characteristics, nearly one-fifth (21.3%) of participants reported they were alcohol drinkers. Whereas, 75 (19%) of respondents chew khat. Moreover, fifty-three (13.5%) of participants conveyed they were performing physical exercise and smoking cigarettes (**Table 1**).

**Table 1:**
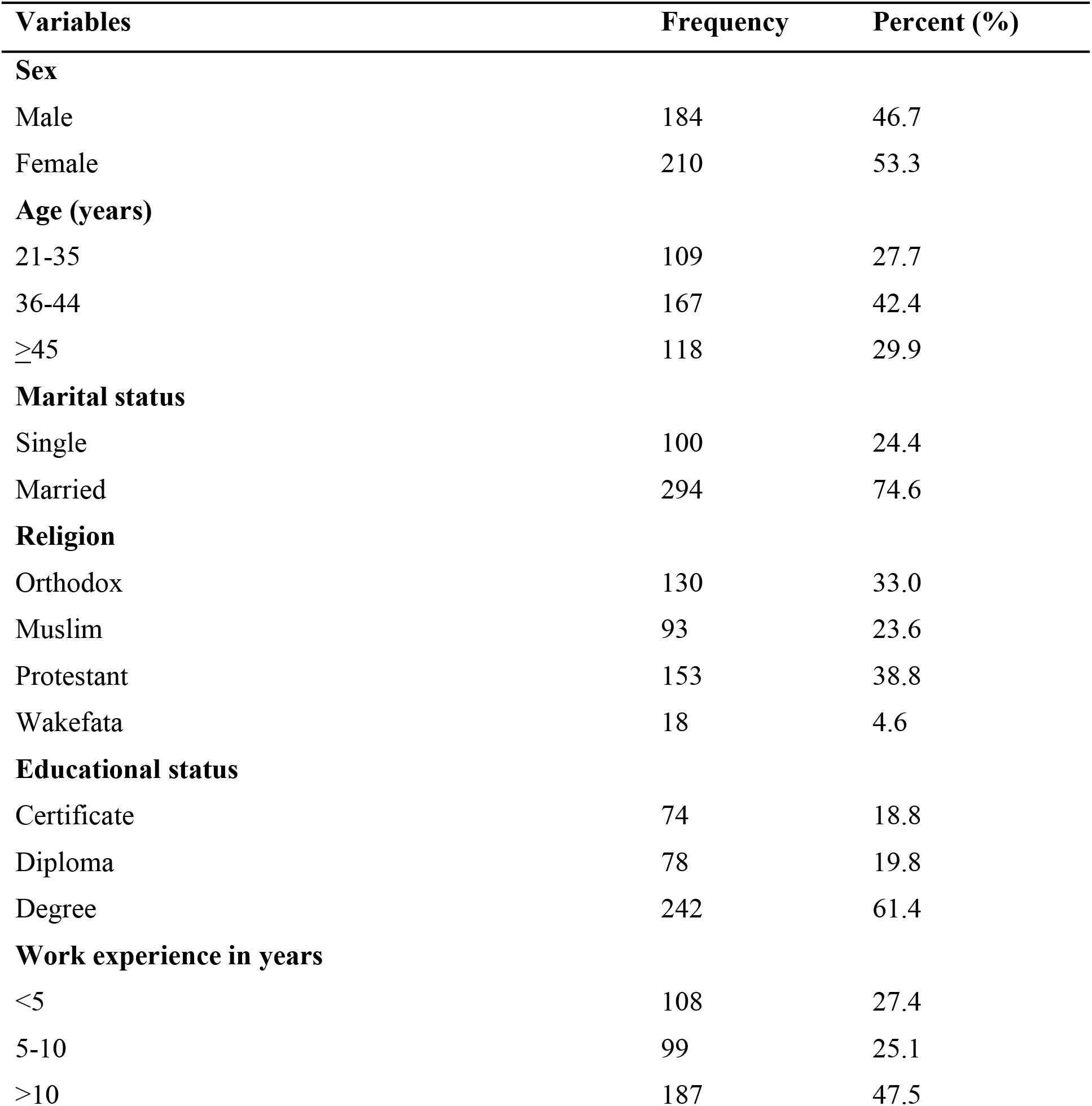
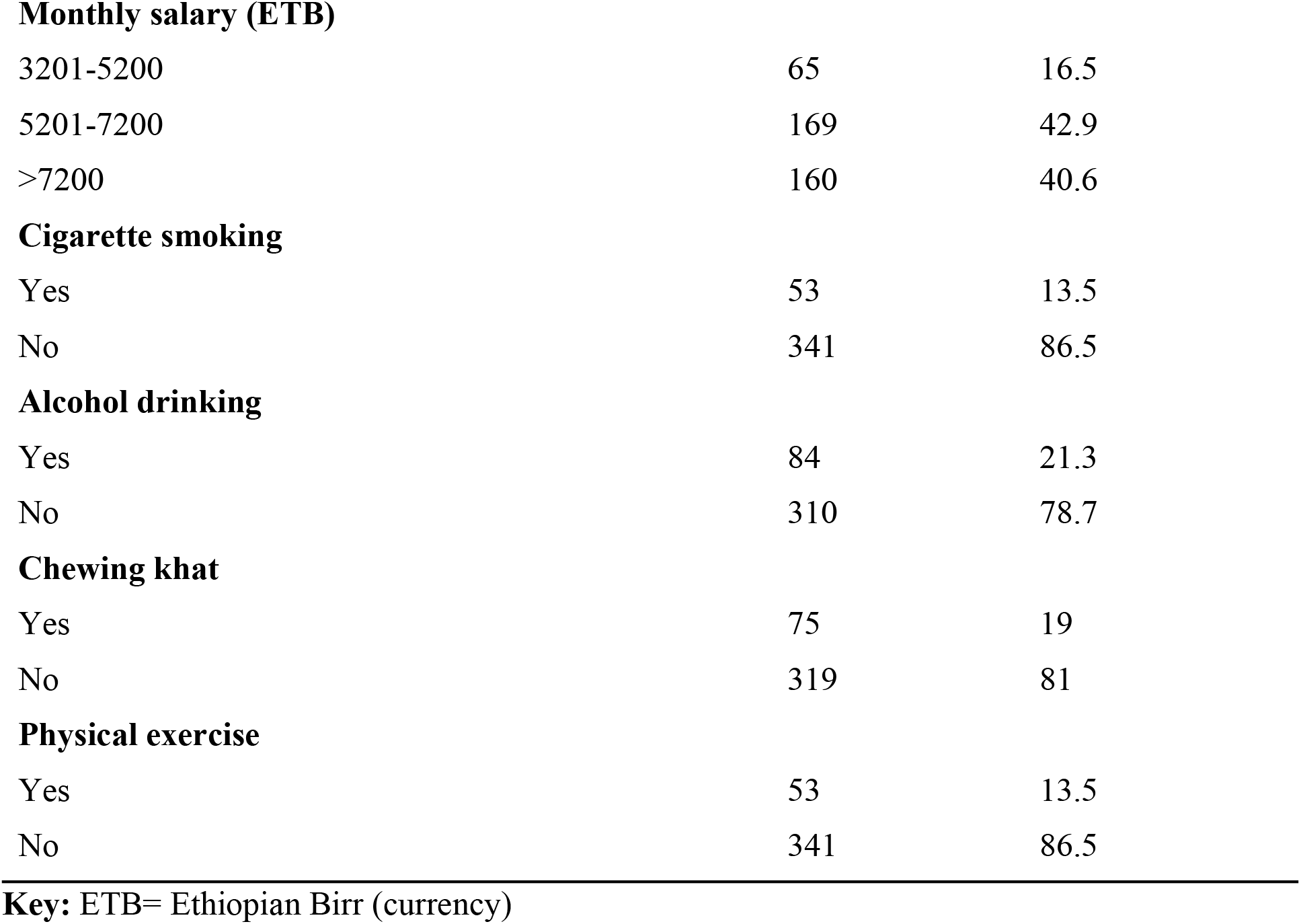
Socio-demographic and behavioral characteristics of study participants among government employees in Nekemte town, Ethiopia, 2021 (N=394).

| Variables | Frequency | Percent (%) |
| --- | --- | --- |
| **Sex** |  |  |
| Male | 184 | 46.7 |
| Female | 210 | 53.3 |
| **Age (years)** |  |  |
| 21-35 | 109 | 27.7 |
| 36-44 | 167 | 42.4 |
| ≥45 | 118 | 29.9 |
| **Marital status** |  |  |
| Single | 100 | 24.4 |
| Married | 294 | 74.6 |
| **Religion** |  |  |
| Orthodox | 130 | 33.0 |
| Muslim | 93 | 23.6 |
| Protestant | 153 | 38.8 |
| Wakefata | 18 | 4.6 |
| **Educational status** |  |  |
| Certificate | 74 | 18.8 |
| Diploma | 78 | 19.8 |
| Degree | 242 | 61.4 |
| **Work experience in years** |  |  |
| <5 | 108 | 27.4 |
| 5-10 | 99 | 25.1 |
| >10 | 187 | 47.5 |
| **Monthly salary (ETB)** |  |  |
| 3201-5200 | 65 | 16.5 |
| 5201-7200 | 169 | 42.9 |
| >7200 | 160 | 40.6 |
| **Cigarette smoking** |  |  |
| Yes | 53 | 13.5 |
| No | 341 | 86.5 |
| **Alcohol drinking** |  |  |
| Yes | 84 | 21.3 |
| No | 310 | 78.7 |
| **Chewing khat** |  |  |
| Yes | 75 | 19 |
| No | 319 | 81 |
| **Physical exercise** |  |  |
| Yes | 53 | 13.5 |
| No | 341 | 86.5 |

### Knowledge, attitude, risk perception, and misconception of COVID-19

One-fourth (25.4%) of participants had poor knowledge about COVID-19, while sixty (15.2%) of employees had poor knowledge of COVID-19 prevention measures. Regarding participants’ attitudes, 223 (56.6%) and 184 (46.7%) of respondents had an unfavorable attitude towards COVID-19 and its preventive measures, respectively. Moreover, a majority (58.1%) of participants were a low-risk perception regarding COVID-19, whereas 175 (44.4%) of respondents had high misconceptions toward 2019 coronavirus disease (**Table 2**).

**Table 2:** Knowledge, attitude, risk perception, and misconception of the study participants about COVID-19 among government employees in Nekemte town, Ethiopia, 2021 (N=394).

| **Variables (n=394)** | **Frequency** | **Percent (%)** |
| --- | --- | --- |
| **Knowledge about COVID-19** |  |  |
| Poor knowledge | 100 | 25.4 |
| Good knowledge | 294 | 74.6 |
| **Knowledge of COVID-19 prevention measures** |  |  |
| Poor knowledge | 60 | 15.2 |
| Good knowledge | 334 | 84.8 |
| **Attitude towards Covid-19 virus** |  |  |
| Unfavorable attitude | 223 | 56.6 |
| Favorable attitude | 171 | 43.4 |
| **Attitude towards prevention measures of COVID-19** |  |  |
| Unfavorable attitude | 184 | 46.7 |
| Favorable attitude | 210 | 53.3 |
| **Risk perception of COVID-19** |  |  |
| High risk | 165 | 41.9 |
| Low risk | 229 | 58.1 |
| **Misconception of COVID-19** |  |  |
| High misconception | 175 | 44.4 |
| Low misconception | 219 | 55.6 |

### Compliance of Personal COVID-19 preventive measures

The finding of this study revealed that the overall utilization of employees’ recommended preventive measures towards COVID-19 was 22.3% (95% CI = 18.5, 26.6).

### Factors associated with Compliance of Personal COVID-19 preventive measures

In the bivariable logistic regression analysis, sex, marital status, physical exercise, attitude towards COVID-19 and its preventive measure, risk perception of COVID-19, and misconception of COVID-19 were the factors associated with utilization of COVID-19 preventive measures. However, after controlling for confounding variables in the multivariable binary logistic regression analysis, only sex, attitude towards COVID-19 prevention measures, risk perception and misconception of COVID-19 remained to have a significant association with compliance of Personal COVID-19 preventive measures. Female workers were almost 3 times more likely to comply COVID-19 personal preventive measures compared with males [AOR = 2.80, 95% CI (1.10, 7.12)], The probability of utilizing Personal COVID-19 preventive measures was 13.73 times greater in employees who had a favorable attitude towards COVID-19 prevention measures compared to those who had unfavorable about it [AOR = 13.73, 95% CI (4.85, 38.83)]. Moreover, the odds of utilizing preventive measures were 2.24 times more likely among employees who had a high-risk perception of COVID-19 than among those who had low-risk perception [AOR=2.34; 95% CI (1.24, 4.41)]. Thus, respondents who had a low misconception of COVID-19 were about 4 times more likely to utilize those measures compared to those who had a high misconception about coronavirus counterpart [AOR = 3.92, 95% CI (1.45, 10.62)] (**Table 3**).

**Table 3:** Logistic regression analysis of factors associated with compliance towards personal COVID-19 preventive measures among Nekemte town government employees, 2021.

| Variables (n=394) | Compliance with PPMs |  | COR with 95% CI | AOR with 95% CI |
| --- | --- | --- | --- | --- |
|  | high | low |  |  |
| Sex |  |  |  |  |
| Male | 19 | 165 | 1 | 1 |
| Female | 69 | 141 | 4.25 (2.44-7.41) | 2.80 (1.10-7.12)* |
| Marital status |  |  |  |  |
| Single | 13 | 87 | 1 | 1 |
| Married | 75 | 219 | 2.29 (1.21-4.34) | 0.61 (0.22-1.74) |
| Physical exercise |  |  |  |  |
| Yes | 8 | 45 | 1 | 1 |
| No | 80 | 261 | 1.72 (0.78-3.81) | 0.59 (0.19-1.81) |
| Attitude towards COVID-19 virus |  |  |  |  |
| Unfavorable attitude | 28 | 195 | 1 | 1 |
| Favorable attitude | 60 | 111 | 3.76 (2.27-6.24) | 0.76 (0.29-1.99) |
| **Attitude towards prevention measures of COVID-19** |  |  |  |  |
| Unfavorable attitude | 9 | 175 | 1 | 1 |
| Favorable attitude | 79 | 131 | 11.73 (5.68-24.23) | 13.73(4.85-38.83)* |
| **Risk perception of COVID-19** |  |  |  |  |
| High risk | 67 | 152 | 3.23 (1.89-5.54) | 2.34 (1.24-4.41)* |
| Low risk | 21 | 154 | 1 | 1 |
| **Misconception of COVID-19** |  |  |  |  |
| High misconception | 12 | 153 | 1 | 1 |
| Low misconception | 76 | 153 | 6.33 (3.31-12.18) | 3.92 (1.45-10.62)* |

**Keys:** 1=reference category AOR=adjusted odds ratio, CI= confidence interval, COR=crudes odds ratio, \*=significant at $p < 0.05$ in multivariable logistic regression analysis, Hosmer and Lemeshow test $p = 0.993$ .

## Discussions

As the lockdowns released in Ethiopia to resume office work, implementation of personal COVID-19 prevention measures in offices is vital in containing the pandemic. We aimed to assess the level of compliance with COVID-19 personal preventive measures and associated factors among a sample of government employees in Nekemte town, western Ethiopia. Accordingly, the analysis showed the high level of compliance with COVID-19 personal preventive measures was stood 22.3% in last one month. The study also revealed sex, attitude, risk perception, and misconception on COVID-19 preventive measures were the factors associated with the compliance of COVID-19 PPMs.

The result of our study was lower than other studies done in Ethiopia (49%) and other regions of the world, such as Nepal (37.1 %), China (53.7%)[40] and (96.8%)[25], Vietnam(75.8%)[41], Japan (90%)[42], Pakistan (46.7%)[43] and Iran (71%) [44]. On the other hand, the current report was higher than a study finding in Thailand (17%)[45]. The disparity could be attributed to differences in study design, sample size, data collection tools and procedures (e.g., web-based, face-to-face, type of tool), study time, and variable operationalization. On top of this, dissimilarities in socio-demographic characteristics, access to protective equipment and comfort, levels of media utilization, individual differences, job satisfaction, position, workplace policy, economic and healthcare system may all have an impact on the level of compliance with personal COVID-19 preventive measures. Furthermore, an Ethiopian study found a high level of reluctance to accept the recommended preventive measures, which is a major issue throughout Ethiopia, including the current study area [46].

In this study, sex of the study participants was found to be significantly associated with level of compliance of COVID-19 PPMs. This findings was supported by previous studies[47], [48], [49] [50]. This could be justified as, female are more conscientious often than males in doing things correctly, likewise, a number of studies that explored the gender effects on individual response to COVID-19 consistently reported that females are more likely to take COVID-19 seriously and vigilantly, and thus are more likely to comply with COVID-19 measures, which may be true for the current study populations[24, 51-53]. Furthermore, sex has been linked to compliance with COVID-19 preventive measures, with men being less likely than women to wash their hands, wear a mask, or engage in social distancing[54]. This could be related to information representing men’s trend to underestimate the brutality of the pandemic[55, 56].

The present study publicized, having a favorable attitudes toward COVID-19 prevention measures leads to greater compliance with personal COVID-19 prevention measures than an unfavorable attitude. This result is consistent with prior research findings [57-59]. This might be explained as government employees with favorable/positive attitudes are more watchful and responsible for implementing preventive measures. Moreover, employees with favorable/positive attitudes comprehend the benefits of prevention, the mode of transmission, and the hazard if it is not prevented properly[60].

The odds of complying with personal COVID-19 preventive measures were two times more likely among employees who had a high-risk perception of COVID-19 than among those who had low-risk perception. This is supported by other scholars[61-65] Undeniably, risk perception is a complex process greatly influenced by many factors including, individuals’ beliefs and perceptions, socio-cultural system, environmental and political conditions, geographic locations, contextual factors, and individual daily experiences[66, 67]. In light of this, high risk perception towards COVID-19 infection could ultimately affect motivation and performance related to their compliance of COVID-19 personal preventive measures [68]

Similarly, respondents who had a low misconception of COVID-19 were about four times more likely to utilize those measures compared to those who had a high misconception about corona virus counterpart. Similar findings were reported from existing literatures [39, 69, 70]. This could be attributed by the fact that the current study was carried out after the disease had spread and public awareness had risen somewhat. These factors have been very important in lowering misconceptions and increasing the implementation of COVID-19 prevention strategies. In addition, it’s crucial to examine the vast and quick spread of numerous conspiracy and false information regarding the ongoing pandemic [71].

In the current study, the following limitations should be considered while interpretations. First, due to the cross-sectional nature of the study design, it is not able to show the temporal relationship between the outcome variable and explanatory variables. Second, we only included government employees who works in offices, so generalization should be made cautiously to individuals working in other types of workplaces. Third, the compliance of COVID-19 personal preventive measures was self-reported by the government employees, which might have introduced recall bias. However, we limited the time to one month, which may lessen remembering issues. Despite these shortcomings, we believe the report strongly depicts government employees’ compliance with COVID-19 personal preventive measures and associated factors.

## Conclusions

In conclusion, little proportion of sampled study participants complied with COVID -19 personal preventive measures. Sex, attitudes, risk perception, and misconception have all been identified as significant risk factors. Actions are needed to strengthen COVID -19 personal preventive measures among government employees to maintain COVID -19 control following work resumption. In the future, it’s vital to work on government employees’ attitudes and perceptions in order to improve compliance. Compliance with advised preventive behaviors has to be improved for employees returning to work during the post-COVID-19 pandemic period.

## Data Availability

All relevant data are within the manuscript and its Supporting Information files.

## Abbreviations

AOR: Adjusted Odds Ratio
CI: Confidence Interval
ETB: Ethiopia Birr
COVID-19: Corona virus disease 19
CPPMs: compliance with COVID -19 personal preventive measures
COR: Crude Odds Ratio
OR: Odds Ratio
SD: Standard Deviation
STATA: Statistics and Data
WHO: World Health Organization

## Acknowledgements

We are very thankful to Nekemte town city administration for approving and providing the necessary preliminary information during this study. We would like to acknowledge the University of Gondar, College of Medicine and Health Sciences and Comprehensive Specialized Hospital, Institute of Public Health for providing ethical clearance. The authors are also very much grateful for all data collectors, supervisors, and the study participants.

